# Plasma microbial cell-free DNA sequencing for the diagnosis and surveillance of fever following hematopoietic stem cell transplant

**DOI:** 10.1101/2023.05.29.23290646

**Authors:** Monica Fung, Nicholas Degner, Srey Sam, Hayley Cleveland, Justin Teraoka, Shraddha Pandey, Dayana Shariff, Timothy Blauwkamp, Peter Chin-Hong

**Affiliations:** University of California San Francisco, San Francisco, California, USA; Karius Inc, Redwood City, California, USA

## Abstract

**Backgroun:** Fever is a common complication in hematopoietic stem-cell transplant recipients, yet an etiology is often not found. Plasma microbial cell-free DNA sequencing may improve diagnosis and management of fever in this population.

**Methods:** This prospective study (ClinicalTrials.gov; NCT02804464) enrolled 70 patients undergoing allogeneic HCT and followed them for one year. Plasma microbial cell-free DNA sequencing was collected within 24 hours of fever and for infection surveillance weekly after HCT through day 63 post-HCT followed by monthly until study end. Plasma mcfDNA sequencing results were compared with all standard microbiological testing results collected within 48 hours of fever onset.

**Results:** Among 53 fevers eligible for analysis, clinical adjudication determined that a probable cause of fever was detected by blood culture in in 7 (13%), by all microbiology in 9 (17%), and by plasma microbial cell-free DNA sequencing in 12 (23%), including 5 in which microbiology was negative, increasing the diagnostic yield by 56%. The positive percent agreement with microbiology was 78% (7/9) and the negative percent agreement was 89% (39/44). Adjudication determined that plasma microbial cell-free DNA sequencing results would have led to a change in antimicrobial management in 55% (7/12) of fevers with detections. Weekly surveillance with plasma microbial cell-free DNA sequencing detected 8 of 15 causative pathogens prior to fever by a mean of 4.8 days (range 1-8 days).

**Conclusions:** Plasma microbial cell-free DNA sequencing shows promise in the surveillance, diagnosis and management of fever in hematopoietic stem cell transplant recipients.

**Article Summary:** Plasma microbial cell-free DNA sequencing increased the diagnostic yield for the infectious etiologies of fever in allogeneic hematopoietic stem-cell recipients by 56% when compared to standard microbiologic testing. Weekly surveillance detected pathogens prior to fever onset.

## INTRODUCTION

Fever is a common complication of hematopoietic cell transplantation (HCT).^1-3^ Given the high risk for morbidity and mortality from infection, fever is often treated with empiric antimicrobial therapy. However, a causative agent is identified by standard microbiology in only approximately a third of patients, and there exist many non-infectious etiologies of fever, leading to potential inappropriate use of antimicrobials.^4-6^ New strategies are needed to improve diagnostic yield, treatment outcomes, and antimicrobial stewardship.

Plasma microbial cell-free DNA (mcfDNA) metagenomic sequencing is a broad, unbiased, non-invasive diagnostic test that may improve diagnosis of infection as compared to current testing strategies reliant upon a battery of microbiologic and molecular tests.^7^ Studies of plasma mcfDNA sequencing have found increased diagnostic yield of culturable, unculturable, atypical, and deep-seated organisms in immunocompromised patients.^8-16^ A prospective study in febrile neutropenia found that plasma mcfDNA sequencing was twice as likely than standard testing to provide a microbiological diagnosis.^17^ However, prospective data on its use in HCT recipients is limited.

Beyond diagnosis, high risk of serious infection in HCT recipients has led to interest in early detection of pathogens prior to the onset of disease through routine surveillance such as with cytomegalovirus (CMV) PCR.^18,19^ Routine surveillance with plasma mcfDNA sequencing may allow early detection and treatment of a broad range of organisms without the limitations of blood culture (fastidious microbes) or PCR (limited panel of organisms detected).

This study examines the performance of the plasma mcfDNA sequencing in the etiologic diagnosis of fever in HCT recipients, its clinical utility in antimicrobial management, and its potential use in routine surveillance for the early detection of infectious causes of fever.

## METHODS

### Study Design and Participants

This prospective, observational study was conducted at University of California, San Francisco Health (UCSF) comparing plasma mcfDNA sequencing as performed by the Karius Test® with the final microbiologic diagnosis by standard microbiologic testing (SMT) for fever (≥38.0°C) in patients undergoing allogeneic HCT (ClinicalTrials.gov Identifier: NCT02804464). The study was approved by the UCSF institutional review board (#15-18026) and eligible patients enrolled after signing an informed consent.

Adults undergoing allogeneic HCT at UCSF were eligible for enrollment prior to conditioning. After enrollment, plasma mcfDNA sequencing was collected in two ways: (1) for diagnosis within 24 hours of any fever with temperature ≥38.0°C and (2) as surveillance for early detection of infection with weekly collections from day 7 post-HCT through day 63 post-HCT followed by monthly collections until 12-months post-HCT. Symptoms, vitals, imaging, and laboratory testing performed within 48 hours of each plasma mcfDNA sequencing collection were recorded. Plasma mcfDNA sequencing results were not made available to the care providers. If a subject had more than one fever in a seven-day period, only the first fever was included in the analysis.

### Plasma Microbial Cell-Free DNA Sequencing Analysis

The Karius Test® was developed and validated in the Karius, Inc. Clinical Laboratory Improvement Amendments-certified/College of American Pathologists-accredited laboratory (Redwood City, California) to detect and quantify mcfDNA in plasma. A detailed description of test methods and validation has been previously described.^7^ An overview of plasma mcfDNA sequencing test procedures and an example clinical report are provided in the Supplementary Appendix. Concentration of plasma microbial cell-free DNA is measured in molecules per microliter (MPM) of plasma. Test results were generated using Karius version 3.11.

### Study Definitions

SMT was defined as the protocol-required minimum diagnostic standard (blood culture) plus any additional diagnostic testing performed during routine care to establish a probable cause of fever. Probable cause of fever was defined as a microbe identified by SMT or plasma mcfDNA sequencing adjudicated as a cause of the febrile event following review of all diagnostic testing and clinical information.

### Primary and Secondary Objectives

The primary study objective was to evaluate the diagnostic performance as measured by the increase in diagnostic yield, positive percent agreement (PPA), and negative percent agreement (NPA) of plasma mcfDNA sequencing as compared to an adjudicated composite of all SMT results available within 48 hours of fever. The secondary objectives were to determine whether real-time availability of plasma mcfDNA sequencing results could have resulted in changes in antimicrobial management (as determined by clinical adjudication) and to evaluate the time to diagnosis for surveillance plasma mcfDNA sequencing performed at prespecified intervals relative to fever onset.

The full analysis data set included all febrile events with data at any timepoint. The modified-intent-to-diagnose (mITD) data set included febrile events with a sample that passed quality control criteria and provided a valid plasma mcfDNA sequencing result, and a valid standard clinical diagnostic result (blood culture). To conservatively estimate agreement with SMT, RNA viruses (RNA is not detected by plasma mcfDNA sequencing) were included in agreement analyses.

SMT was considered positive if it detected at least one organism adjudicated as a probable cause of fever, even if other detected organisms were adjudicated as an unlikely cause of the fever. If SMT detected no organisms or if all organisms detected were adjudicated as an unlikely cause of the fever, it was classified as negative. The same rules applied to plasma mcfDNA sequencing results. For the calculation of PPA and NPA, plasma mcfDNA sequencing results were compared with the clinically adjudicated positive and negative SMT results.

### Surveillance for early detection of infection

For each fever in which a probable cause was detected by plasma mcfDNA sequencing, the preceding plasma mcfDNA sequencing collected as part of surveillance were assessed to determine if the probable cause was detected prior to fever onset and if so, the number of days prior to fever onset it was first detected.

### Endpoint Adjudication Process

Following a standardized review of vitals, symptoms, laboratory data, radiology results, medications, and the electronic medical record, each of 2 infectious diseases specialists (study authors MF and HC) independently classified results from SMT and plasma mcfDNA sequencing as a probable or unlikely cause of the febrile event. Adjudicators were blinded to plasma mcfDNA sequencing results when adjudicating SMT. Once all eligible febrile events had been independently adjudicated, any discrepancies were resolved through a consensus meeting with the two adjudicators. The adjudicators also evaluated the antimicrobial management for each febrile event and determined whether real-time availability of the plasma mcfDNA sequencing results could have changed antimicrobial management through broadening, narrowing or targeting, or confirming patient was on the appropriate regimen.

### Statistical Analysis

Diagnostic performance measures are reported as percent with two-sided Clopper-Pearson exact 95% binomial confidence limits. Continuous variables were compared using a t-test and categorical variables were compared using Fisher’s exact test. All statistical analyses were performed using SAS statistical software, version 9.4.

## RESULTS

### Patient Characteristics and Febrile Events

A total of 70 patients were enrolled from August 2016 to April 2018. The most common indications for HCT were acute myeloid leukemia (44%), acute lymphoblastic leukemia (14%), myelodysplastic syndrome (11%), and chronic lymphocytic leukemia (10%) (Table 1). Of the 70 patients, 16 died (23%) and 6 withdrew (9%) prior to study completion at 12 months.

**Table 1.**
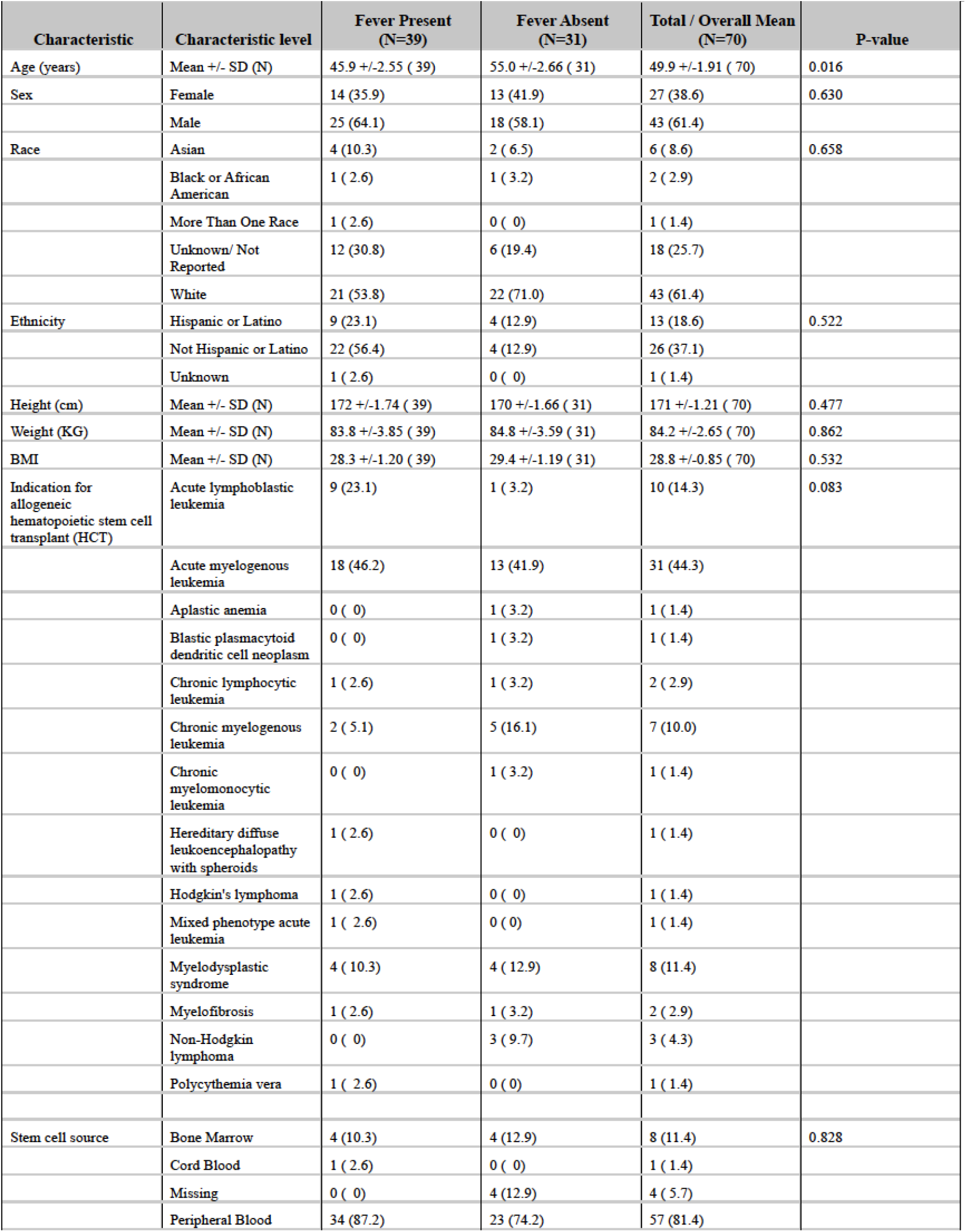
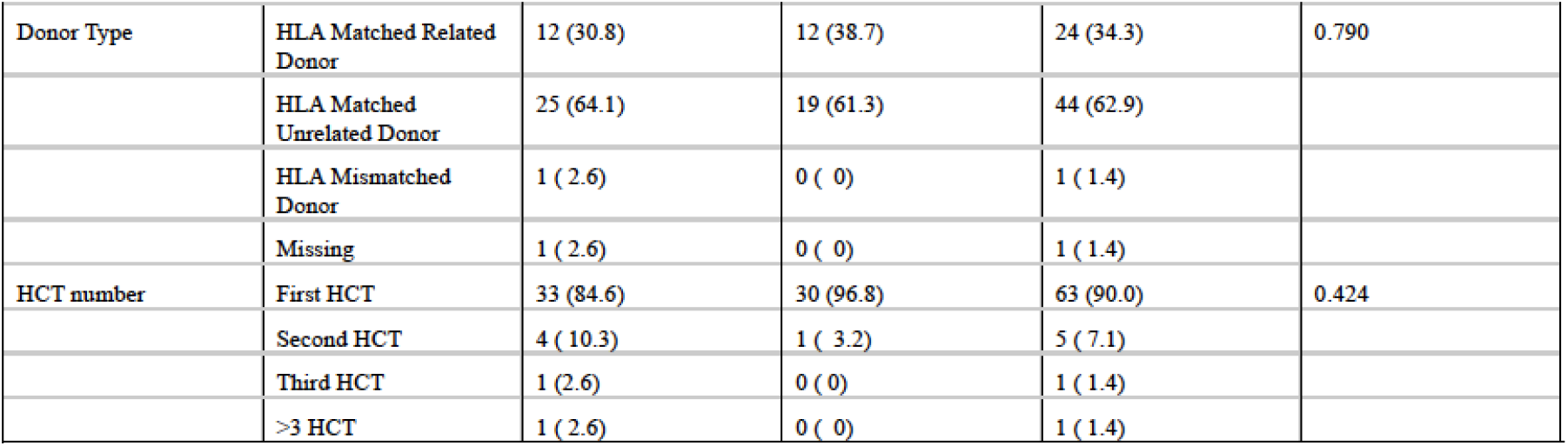
Demographics & Medical History. Patient Demographics and Characteristics.

A total of 65 fevers were captured among 39 patients. The mITD group for analysis included 53 of these 65 fevers, as 12 of the fevers occurred within 7 days of a prior fever in the same subject (Figure 1). The median time from HCT to fever was 10 days (mean 23.6 days; range -9 to 170 days) and the distribution of fevers is shown in Figure 2. The only significant difference in demographics and medical history between those with and without fever was age (mean 45.9 years vs 55.0 years with vs. without fever, Table 1). Of the 53 fevers, 38 (72%) occurred in patients with neutropenia (<1500 absolute neutrophil count [ANC]/microL), 28 (53%) occurred in patients with severe neutropenia (<500 ANC/microL), and 23 (43%) occurred in patients with profound neutropenia (<100 ANC/microL).

**Figure 1.**
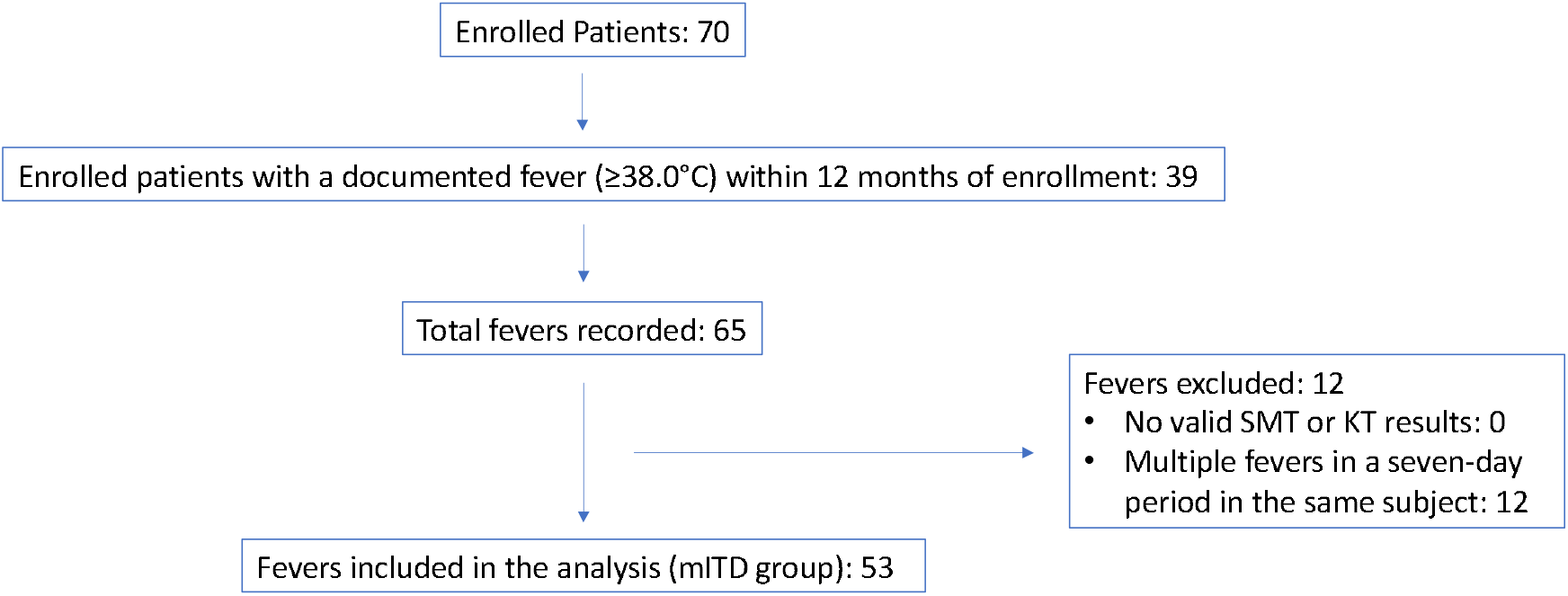
Patient enrollment and fevers included in the analysis. The modified-intent-to-diagnose (mITD) data set included febrile events with a sample that passed quality control criteria, providing a valid KT result, and a valid standard clinical diagnostic result for the same specimen collection time. If a subject had more than one fever (≥38.0C) in a seven-day period, only the first fever was included in the mITD data set.

**Figure 2.**
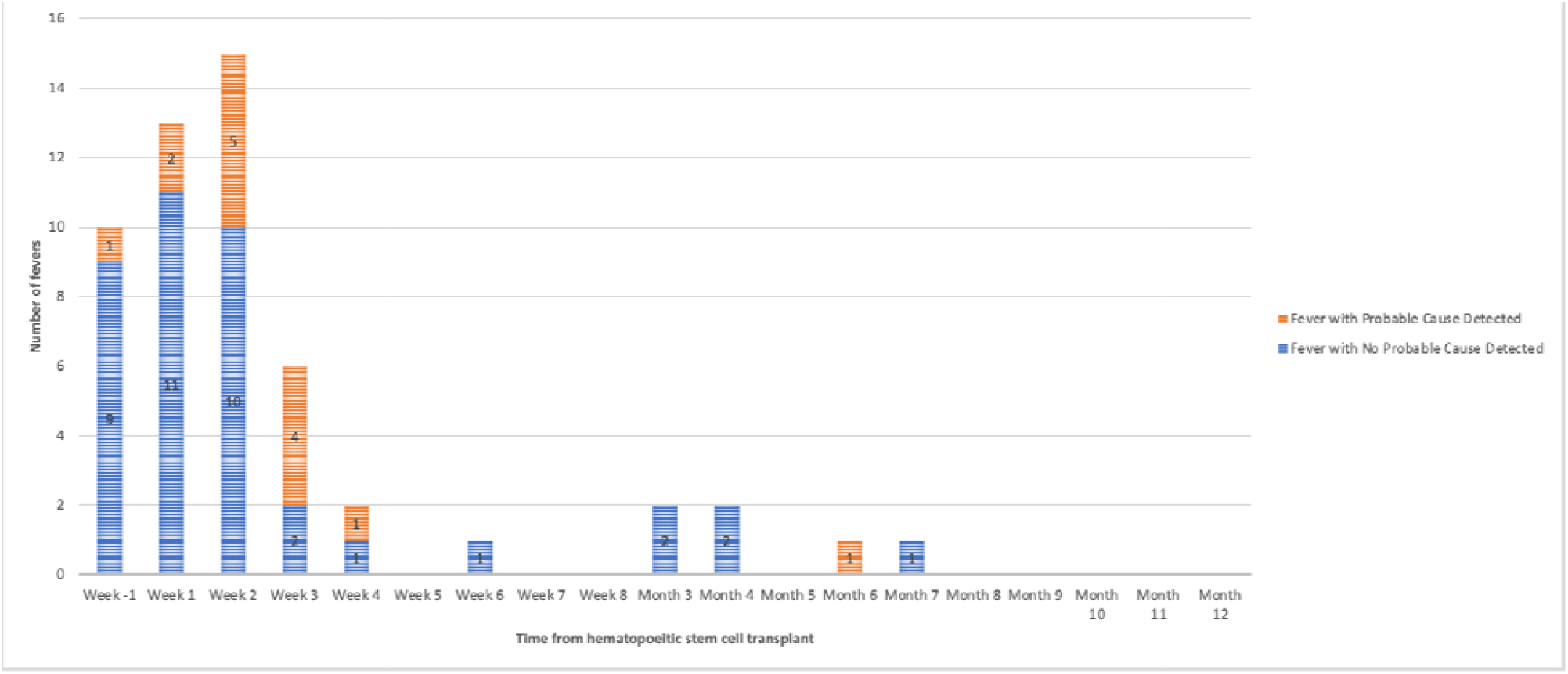
Distribution of fevers in the study relative to date of hematopoietic stem cell transplantation.

### Fever Diagnostic Outcome

At least one adjudicated probable cause of fever was identified by blood culture collected within 48 hours of fever in 7/53 (13%) fevers and by all SMT collected within 48 hours of fever in 9/53 (17%) fevers (Figure 3 and Table 2). Of these, 2 probable causes of fever were identified in 1/9 (11%) fevers and 3 probable causes of fever were identified in 1/9 (11%) fevers. Among 12 probable causes of fever, 8 (67%) bacterial, 2 (17%) DNA viral, 1 (8%) RNA viral, and 1 (8%) fungal pathogens were identified by SMT.

**Table 2.** Fevers with a probable cause identified by standard microbiologic testing or plasma microbial cell-free DNA (mcfDNA) sequencing. HCT: allogeneic hematopoietic stem cell transplant. “-” indicates no probable cause was identified. MPM: molecules per microliter (concentration of plasma microbial cell-free DNA detected). NOS: not otherwise specified.

| Days from HCT | Infectious Syndrome | Probable Causes of Fever Detected by Standard Microbiology Testing | Probable Causes of Fever Detected by Plasma mcfDNA Sequencing (MPM) | Unlikely Causes of Fever Detected by Standard Microbiology Testing | Unlikely Causes of Fever Detected by Plasma mcfDNA Sequencing (MPM) |
| --- | --- | --- | --- | --- | --- |
| -6 days | Neutropenic Fever NOS | - | *Streptococcus mitis* (77) | - | - |
| 4 days | Neutropenic Fever NOS | - | *Gemella haemolysans* (86) | *Enterococcus spp* (Urine) | - |
| 7 days | Neutropenic Fever, Bloodstream Infection | *Escherichia coli*<br>*Staphylococcus epidermidis* | *Staphylococcus epidermidis* (389) | - | - |
| 10 days | Neutropenic Fever, Bloodstream Infection | *Fusobacterium nucleatum* | - | *Staphylococcus epidermidis* (Blood)<br>*Cytomegalovirus* (Blood) | *Staphylococcus epidermidis* (719)<br>*Cytomegalovirus* (261) |
| 10 days | Neutropenic Fever, Upper Respiratory Tract Infection | *Human metapneumovirus* | - | - | - |
| 11 days | Bloodstream Infection | *Streptococcus mitis* | *Streptococcus mitis* (9364) | - | *Cytomegalovirus* (19) |
| 12 days | Neutropenic Fever, Bloodstream Infection | *Enterococcus faecalis* | *Enterococcus faecalis* (590967) | - | *Herpes Simplex Virus-2* (518) |
| 14 days | Neutropenic Fever, Bloodstream Infection, Colitis | *Candida krusei*<br>*Cytomegalovirus*<br>*Enterococcus gallinarum* | *Candida krusei* (218833)<br>*Enterococcus gallinarum* (41331)<br>*Enterococcus faecium* (26452)<br>*Cytomegalovirus* (17860) | Mixed gram positive flora (Endotracheal)<br>*BK polyomavirus* (Blood) | *Staphylococcus epidermidis* (635063)<br>*Staphylococcus haemolyticus* (326179)<br>*BK polyomavirus* (54621) |
| 15 days | Pneumonia | - | *Mycoplasma hominis* (19) | Mixed flora (Urine)<br>*Cytomegalovirus* (Blood) | *Human herpes virus 6B* (555)<br>*Cytomegalovirus* (150)<br>*Bacillus cereus* (109)<br>*BK polyomavirus* (59)<br>*Bacillus thuringiensis* (36) |
| 16 days | Neutropenic Fever, Neutropenic Enterocolitis | - | *Bacteroides vulgatus* (38)<br>*Fusobacterium nucleatum* (33) | - | - |
| 17 days | Neutropenic Fever, Bloodstream Infection | *Enterococcus faecium* | *Enterococcus faecium* (82766) | - | *Herpes Simplex Virus-2* (126) |
| 19 days | Neutropenic Fever NOS | - | *Cytomegalovirus* (6260) | - | *Human herpes virus 6B* (65) |
| 22 days | Neutropenic Fever, Bloodstream Infection | *Staphylococcus aureus* | *Staphylococcus aureus* (4042923) | Mixed gram positive flora (Endotracheal) | - |
| 169 days | Hemorrhagic Cystitis | *BK polyomavirus* | *BK polyomavirus* (82) | - | *Cyberlindnera fabianii* (175) |

**Figure 3.**
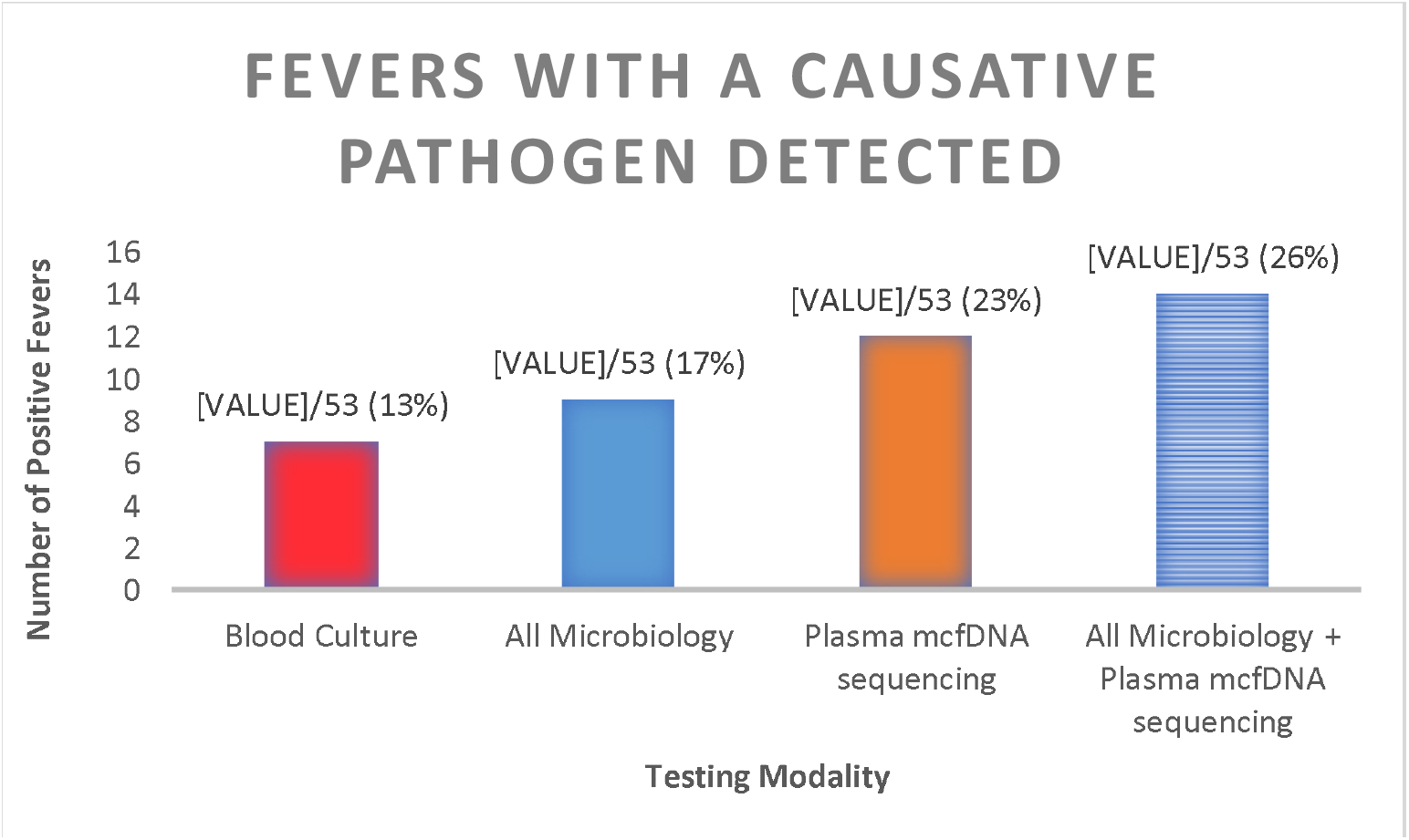
Number of fevers with a probable cause identified as determined by clinical adjudication by testing modality. mcfDNA: microbial cell-free DNA.

Plasma mcfDNA sequencing collected within 24 hours of fever identified an adjudicated probable cause of fever in 12/53 (23%) fevers (Figure 3 and Table 2). Of these, 2 probable causes of fever were identified in 1/12 (8%) and 4 probable causes of fever were identified in 1/12 (8%). Among 16 probable causes of fever, 12 (75%) bacterial, 3 (19%) DNA viral, and 1 (6%) fungal pathogens were identified by plasma mcfDNA sequencing. The combination of SMT and plasma mcfDNA sequencing identified a probable cause of fever in 14/53 (26%) fevers (Figure 3 and Table 2).

### Diagnostic Performance

Plasma mcfDNA sequencing exclusively identified a probable cause of fever in 5/53 (9.4%) fevers, equating to a 9.4% absolute increase in diagnostic yield, and a 56% relative increase in diagnostic yield (increase in fevers with a probable cause detected from 9 to 14/53) (Table 3). Probable causes of fever identified by plasma mcfDNA but not SMT collected within 48 hours of fever onset were *Bacteroides vulgatus, CMV, Enterococcus faecium, Fusobacterium nucleatum, Gemella haemolysans, Mycoplasma hominis*, and *Streptococcus mitis* (Table 2). The CMV was eventually detected by PCR sent 72 hours after fever onset (outside the 48 hour study window).

**Table 3.** Diagnostic Performance. Positive and negative percent agreement between plasma microbial cell-free DNA (mcfDNA) sequencing and microbiology in detecting at least one probable cause of fever.

|  | All microbiology |  |  |  |
| --- | --- | --- | --- | --- |
|  |  | Probable Cause Detected | No Probable Cause Detected | Total |
| **Plasma mcfDNA sequencing** | **Probable Cause Detected** | 7 | 5 | 12 |
|  | **No Probable Cause Detected** | 2 | 39 | 41 |
|  | **Total** | 9 | 44 | 53 |
| **Measure** |  |  |  |  |
| Positive percent agreement | 77.8% (7/9) |  |  |  |
| Negative percent agreement | 88.6% (39/44) |  |  |  |
| Increase in diagnostic yield (absolute) | 9.4 (5/53) |  |  |  |
| Increase in diagnostic yield (relative) | 55.6 (5/9) |  |  |  |

Among 9 fevers with a probable cause identified by SMT, plasma mcfDNA identified at least one of the same pathogens in 7 (PPA 78%, Table 3). Probable causes of fever identified by SMT but not plasma mcfDNA sequencing were *Escherichia coli, Fusobacterium nucleatum*, and Human metapneumovirus (Table 2). Of 44 fevers with no probable cause identified by SMT, plasma mcfDNA did not identify a probable cause of fever in 39 (NPA 89%, Table 3). Of the 7 cases in which both SMT and plasma mcfDNA sequencing found a probable cause of fever, there was complete concordance on the probable causes in 5 of the cases. Of the remaining two, in one plasma mcfDNA sequencing found one additional probable cause (*Enterococcus faecium*), and in the other SMT found one additional probable cause (*Escherichia coli*) (Table 2).

Organisms adjudicated as unlikely causes of fever were detected by SMT in 17 of 53 fevers (32%) and by plasma mcfDNA sequencing in 27 of 53 fevers (51%) (Supplementary Data). The most frequent pathogens adjudicated as unlikely causes of fever detected by plasma mcfDNA sequencing (n>2) were CMV (n=10), *Staphylococcus epidermidis* (n=6), BK *polyoma virus* (n=5), *Enterococcus faecium* (n=3), *Human herpesvirus-6* (n=3), *Herpes simplex virus-1* (n=3) (Supplementary Data).

### Potential Clinical Impact of Plasma mcfDNA Sequencing

Of 12 fevers in which a probable cause was identified by plasma mcfDNA sequencing, adjudication determined that had the plasma mcfDNA sequencing results been known in real-time, it would have led to a narrowing or targeting of antimicrobial coverage in 6 (50%), confirmed that the patient was on the appropriate antimicrobial regimen in 5 (42%), and led to broadening of coverage in 1 (8%) (Table 4). In total 7 (58%) would have had a change in their antimicrobial regimen.

**Table 4.** Clinical impact of plasma microbial cell-free DNA (mcfDNA) had the results been available in real time as determined by clinical adjudication in fevers in which plasma mcfDNA sequencing detected the probable cause of fever.

Clinical impact of plasma mcfDNA sequencing detections
| Outcome | Proportion (%) |
| --- | --- |
| Narrowing or targeting of antimicrobial coverage | 6/12 (50.0) |
| Confirmed patient is on appropriate antimicrobial regimen | 5/12 (41.7) |
| Broadening of antimicrobial coverage | 1/12 (8.3) |

### Routine surveillance with plasma mcfDNA sequencing

Among the 12 fevers in which a probable cause was identified by plasma mcfDNA sequencing collected at time of fever, 11 had surveillance collected prior to the onset of fever. Of those 11, routine surveillance detected a probable cause prior to fever onset in 7 (64%) by a mean of 4 days (median 3.5 days; range 1-8 days) (Table 5 and Figure 4). On a per organism basis, plasma mcfDNA sequencing identified 16 pathogens, of which 15 had prior surveillance KT testing. Of these 15 pathogens, 8 (53%) were identified prior to fever onset by plasma mcfDNA sequencing surveillance by a mean of 4.8 days (median 6 days; range 1-8 days) (Table 5 and Figure 4). The mean MPM of mcfDNA at time of early detection by surveillance was 10,617 (median 2,094 MPM; range 61 to 64,633 MPM) compared to a mean MPM of mcfDNA of 601,072 at time of fever (median 33,892 MPM; range 19 to 4,042,923 MPM; p<0.001) (Table 5).

**Table 5.** Plasma microbial cell-free DNA (mcfDNA) surveillance. After enrollment patients received weekly collection of plasma microbial cell-free DNA sequencing for surveillance until day 63 after hematopoietic stem cell transplant at which time it was collected monthly until 12 months after enrollment. Listed here are the nearest surveillance results for the 12 fevers in which a probable cause was identified by plasma mcfDNA sequencing.

| Probable Cause of Fever Detected by Karius Test at Time of Fever | Days Prior to Fever Onset of Most Recent Karius Test Surveillance | Molecules per microliter of pathogen plasma cell-free DNA at surveillance collection prior to fever onset | Molecules per microliter of pathogen plasma cell-free DNA at time of fever |
| --- | --- | --- | --- |
| *Mycoplasma hominis* | 1 | 61 | 19 |
| *Staphylococcus aureus* | 1 | 11,005 | 4,042,923 |
| *Enterococcus faecium* | 2 | 64,633 | 82,766 |
| *Bacteroides vulgatus* | 3 | Not detected | 38 |
| *Fusobacterium nucleatum* | 3 | Not detected | 33 |
| *Gemella haemolysans* | 3 | Not detected | 86 |
| *Streptococcus mitis* | 4 | Not detected | 9364 |
| *Enterococcus faecalis* | 5 | 1612 | 590,967 |
| *Candida krusei* | 7 | Not detected | 218,833 |
| *Cytomegalovirus* | 7 | 110 | 17,860 |
| *Enterococcus faecium* | 7 | 4,583 | 26,452 |
| *Enterococcus gallinarum* | 7 | 2,576 | 41,331 |
| *Cytomegalovirus* | 8 | 355 | 6,260 |
| *Staphylococcus epidermidis* | 8 | Not detected | 389 |
| *BK polyomavirus* | 10 | Not detected | 82 |

**Figure 4.**
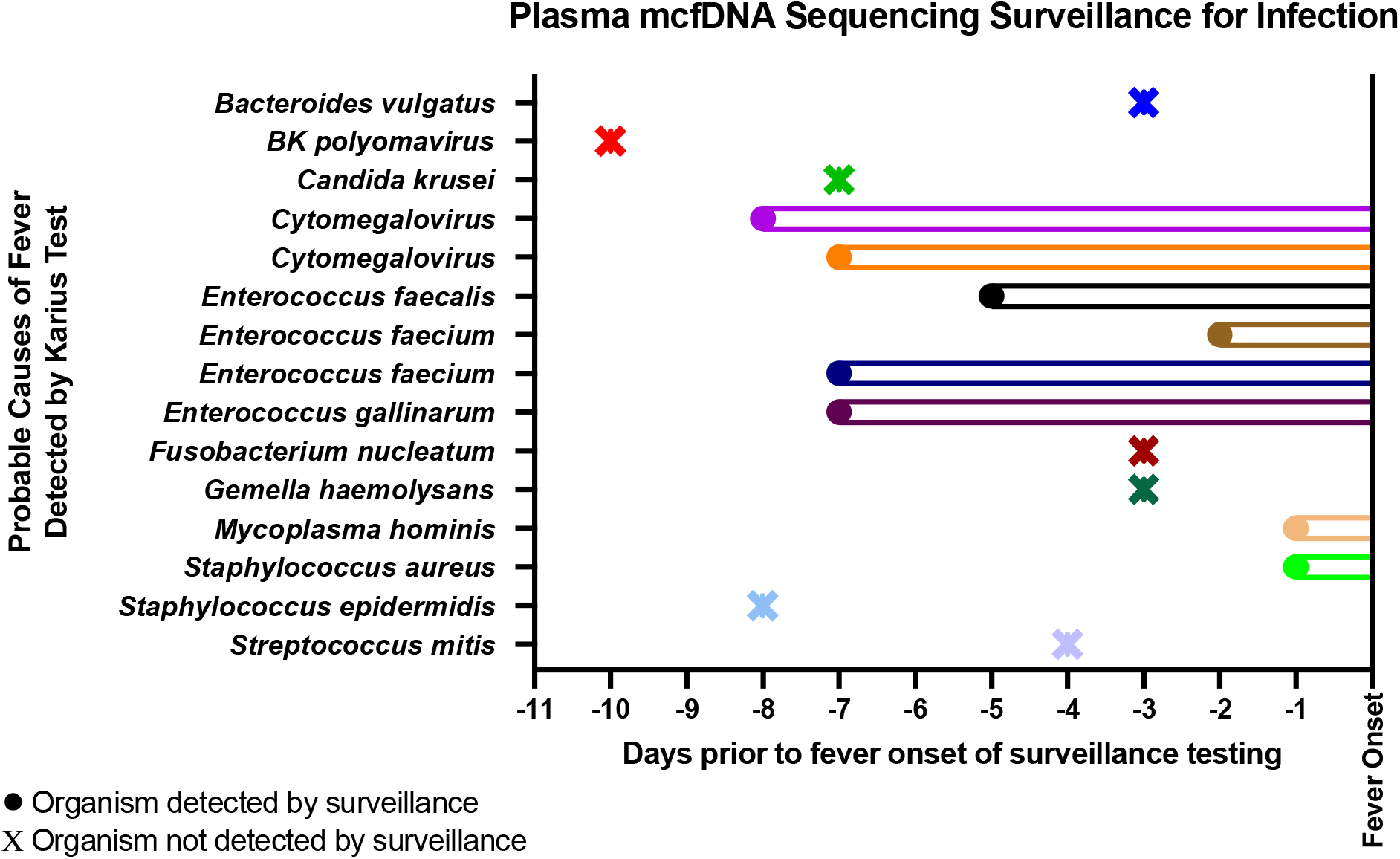
Plasma microbial cell-free DNA (mcfDNA) surveillance for infection. After enrollment patients received weekly collection of plasma mcfDNA sequencing for surveillance until day 63 after hematopoietic stem cell transplant at which time it was collected monthly until 12 months after enrollment. Displayed here is the result of the most recent surveillance plasma mcfDNA sequencing prior to fever onset with a circle and solid line indicating detection and the X symbol with no line indicating the organism was not detected. Of the 15 pathogens detected as probable causes of fever, plasma mcfDNA sequencing surveillance detected 8 prior to fever onset and did not detect 7 prior to fever onset.

## DISCUSSION

In this prospective study in HCT-recipients, adding plasma mcfDNA sequencing to SMT increased the number of fevers with an infectious cause detected from 17% to 26%, increasing the overall diagnostic yield by 56%. Importantly, plasma mcfDNA sequencing was able to identify fastidious bacteria (*Mycoplasma hominis*) and anaerobes (*Fusobacterium, Bacteroides*) as well as viruses requiring dedicated PCRs (CMV, BK virus), emphasizing several strengths of plasma mcfDNA sequencing, namely overcoming the limitations of culture-based methods and unbiased testing. Furthermore, the majority of plasma mcfDNA sequencing diagnoses were adjudicated as likely to lead to treatment changes.

An infectious etiology was detected by blood culture in 13% of fevers, by all SMT in 17% of fevers, and by plasma mcfDNA sequencing 23% of fevers in this study. While low, this finding is comparable with studies in febrile neutropenia reporting yields of 10-20% for blood culture and 20-30% for all SMT.^4^ In this study, at the time of fever 72% of patients had neutropenia (<1500 ANC/microL) and 53% had severe neutropenia (<500 ANC/microL). Additionally, many patients were on prophylactic or treatment antimicrobials at the onset of fever. Previous studies have shown that plasma mcfDNA DNA remains detectable longer than conventional blood culture in patients with prior antibiotic therapy, which may partially explain the increased diagnostic yield seen with plasma mcfDNA sequencing.^20,21^ The low diagnostic yield also highlights the numerous potential non-infectious causes of fever in this population, including drug fever, engraftment and graft-versus-host disease (GVHD).^1,2^ Indeed, 23% of fevers with no probable cause detected occurred within a week of engraftment and 72% occurred in patients with GVHD. Further studies on the clinical utility of plasma mcfDNA organism quantification and integration of host-response-based gene signatures may help augment the diagnostic performance and clinical management.^22,23^

Overall, plasma mcfDNA sequencing demonstrated high concordance with probable detections by SMT with a PPA of 78% and a NPA of 89%. Of the nine fevers with a probable cause identified by SMT, plasma mcfDNA sequencing did not detect a probable cause in two, and in a third it only detected one of two organisms. Of these three organisms, two were bacteria (*Fusobacterium nucleatum and Escherichia coli*) grown in blood culture in which mcfDNA for this organism was detected in the sample, however they did not meet the established statistical threshold for reporting, and the third was a respiratory viral panel positive for human metapneumovirus, an RNA virus inherently undetectable by DNA sequencing.

Most causative pathogens detected were bacteria (74%), followed by viruses (21%), and a single fungus (5%), *Candida krusei*. Half of the infectious syndromes in which these pathogens were detected were blood stream infections followed by neutropenic fever without localizing symptoms (21%). The Organ Transplant Infection Project similarly found that bacteremia was the most common infection, however it found higher rates of invasive fungal infections.^3^ These differences are consistent with the fact 86% of fevers with a causative pathogen identified in this study occurred in the pre-engraftment period in the 30 days following HCT, as fever during this period is usually caused by translocation of endogenous bacteria or *Candida* species into the bloodstream.^1,2^ Only 14% of fevers were in the post-engraftment period, reflecting the low rate of invasive fungal infections, potentially due to standard antifungal prophylaxis at our center.

Beyond increased diagnostic yield, plasma mcfDNA sequencing may assist with antimicrobial stewardship. Specifically, had plasma mcfDNA sequencing results been known in real-time, it would have led to a change in antimicrobials in 58% of cases where it detected a probable cause of fever. Given the burden of multidrug resistant organisms in the HCT population, it is critical to balance antimicrobial stewardship to reduce development of drug resistance and antimicrobial toxicity with appropriate therapy to reduce adverse infectious outcomes.^6^

Weekly routine scheduled surveillance with the plasma mcfDNA sequencing detected 53% of causative pathogens prior to fever onset by a mean of 4.8 days (range 1-8 days) and showed a clear increase in mcfDNA concentration from first detection to fever onset. This evidence builds upon a study of plasma mcfDNA sequencing for predicting bloodstream infections in pediatric patients with relapsed or refractory cancer in which in the 3 days before culture positivity the predictive sensitivity of plasma mcfDNA was 75%.^10^ Given the severity of infections in HCT-recipients and the importance of early administration of antimicrobial therapy, this is a potentially promising application of the test in this population. However, given organisms adjudicated as unlikely causes of fever were also identified in separate instances as probable causes of fever, interpretation of surveillance plasma mcfDNA sequencing results in asymptomatic HCT recipients requires significant further study.

While our study demonstrates the promise of plasma mcfDNA sequencing in HCT recipients, several limitations exist. The sample size of 70 patients with 53 fevers limited power to detect significant differences in diagnostic performance. Another limitation is that the study centered around capturing febrile events, which is likely easier in the initial hospitalization after HCT than after discharge to the outpatient setting, biasing the fevers included in the study towards the pre-engraftment period. Further, SMT tests collected beyond 48 hours after the onset of fever were not included. Next, it was a single center study and diagnostic and antimicrobial prophylaxis practices among other factors may have differed from other centers limiting generalizability. Finally, it is also important to point out the need for expert clinical interpretation of all organisms detected by plasma mcfDNA sequencing.

In summary, HCT recipients are at high risk of infection-related morbidity and mortality, and improved diagnostics that can capture the large range of potential pathogens in a timely fashion are needed. Plasma mcfDNA sequencing demonstrated high concordance with SMT and added to diagnostic yield with the potential clinical utility of improving appropriate use of antimicrobials.

## Data Availability

All data produced in the present study are available upon reasonable request to the authors.

## Acknowledgements

M.F.: study design, data collection, clinical review, manuscript writing and revision. N.D.: study design, data analysis, manuscript writing and revision. S.S.: study design, data collection, data analysis. H.C.: data collection, clinical review. J.T.: data collection. S.P.: data collection. D.S.: data collection. T.B.: study design, manuscript revision. P.C.: study design.

## Potential conflicts of interest

M.F.: the work associated with this study was funded by Karius, Inc. N.D.: Employed at Karius, Inc. during the conduct of this study. S.S.: Employed at Karius, Inc. during the conduct of this study. H.C.: the work associated with this study was funded by Karius, Inc. J.T.: the work associated with this study was funded by Karius, Inc. S.P.: the work associated with this study was funded by Karius, Inc. D.S.: the work associated with this study was funded by Karius, Inc. T.B.: Employed at Karius, Inc. during the conduct of this study. P.C.: the work associated with this study was funded by Karius, Inc.

*Data available in supplementary material*.

