## Supplemental Data for "Plasma microbial cell-free DNA sequencing for the diagnosis and surveillance of fever following hematopoietic stem cell transplant"

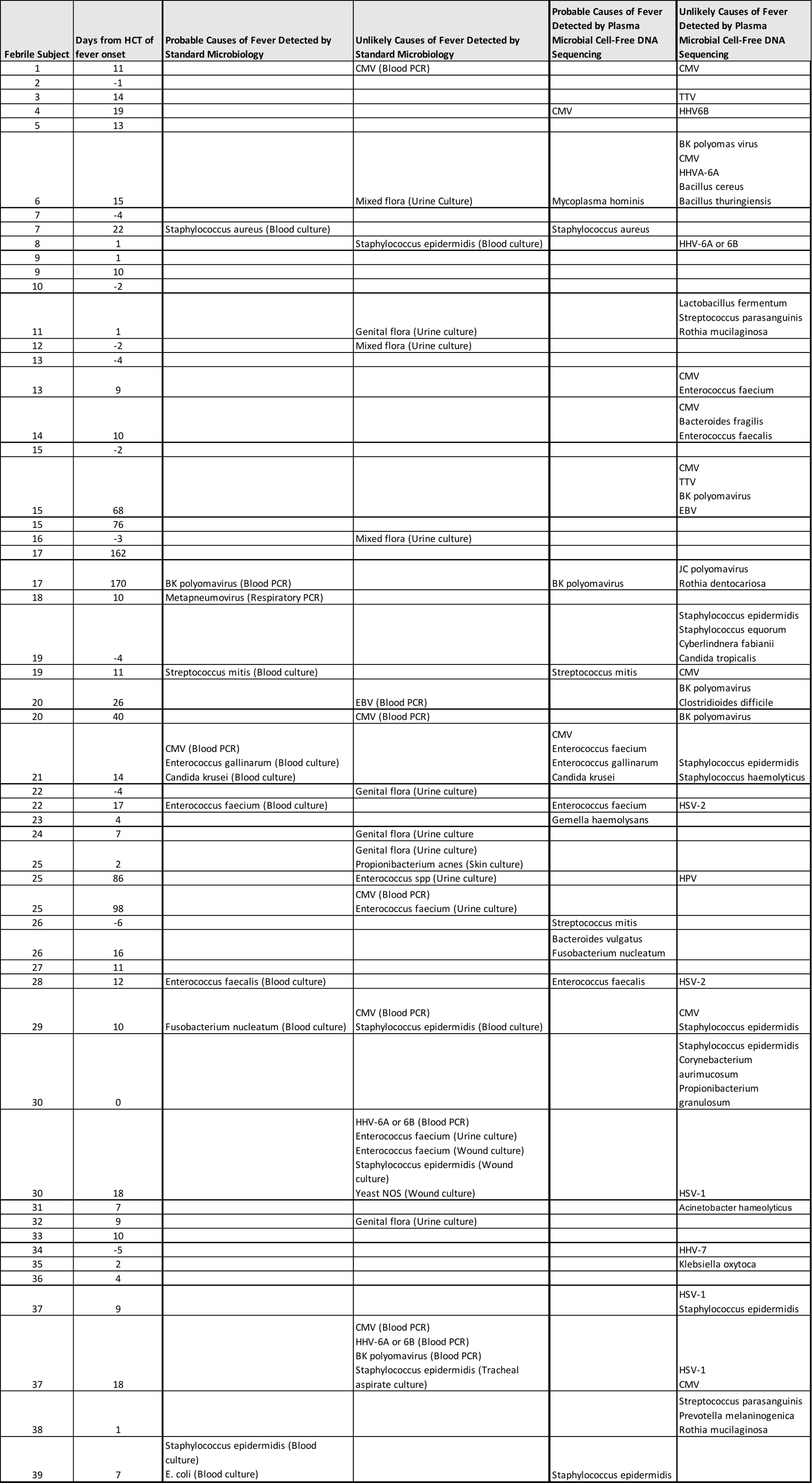

**Supplemental Data.** Clinical adjudication results for each fever for each organism detected by standard microbiology performed with 48 hours of fever or plasma microbial cell-free DNA sequencing performed within 24 hours of fever.
