## Supplemental Appendix for "Plasma microbial cell-free DNA sequencing for the diagnosis and surveillance of fever following hematopoietic stem cell transplant"

Plasma Microbial Cell-Free DNA Sequencing Test Procedure

The Karius Test® was developed and validated in the Karius Clinical Laboratory Improvements Amendments-certified/College of American Pathologists-accredited/New York State Department of Health-approved laboratory (Redwood City, California) to detect and quantify mcfDNA in plasma. After collection, blood samples were processed, and plasma separated and shipped to Karius following the Karius Specimen Collection & Preparation Instructions protocol (<https://kariusdx.com/karius-test/karius-test-process#specimen-collection>). Following mcfDNA extraction, enriched library preparation, sequencing (Illumina NextSeq550), discarding of human sequences, and alignment of the remaining sequences to a curated database of reference genomic sequences, test results were generated using the Karius version 3.11 analytical pipeline. This bioinformatic pipeline was designed to detect and quantify 1,563 microbes across bacteria, DNA viruses, fungi, and other eukaryotes. Plasma mcfDNA of microorganisms that are determined to be significantly higher than real-time background control specimens were reported and quantified in molecules per microliter, which is equivalent to the number of sequencing reads per microliter of plasma.

Example Plasma Microbial Cell-Free DNA Sequencing Test Report

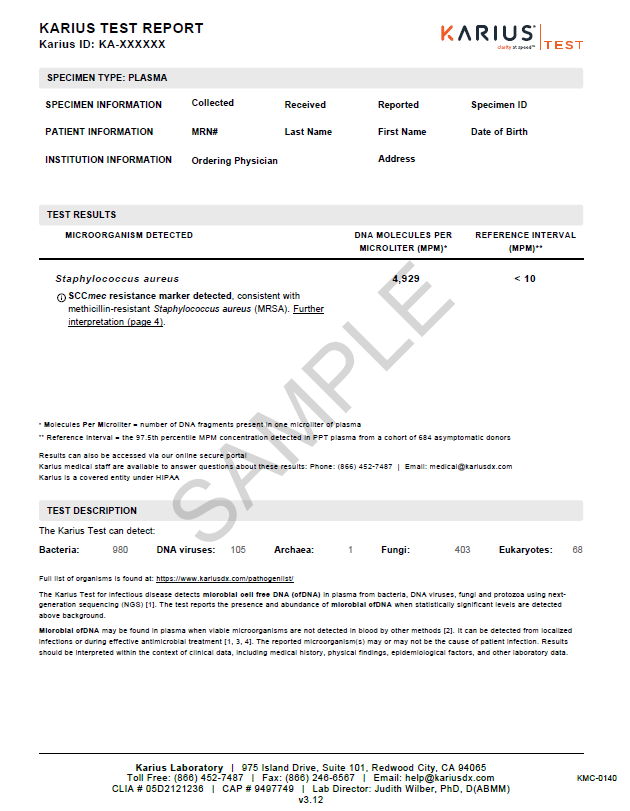

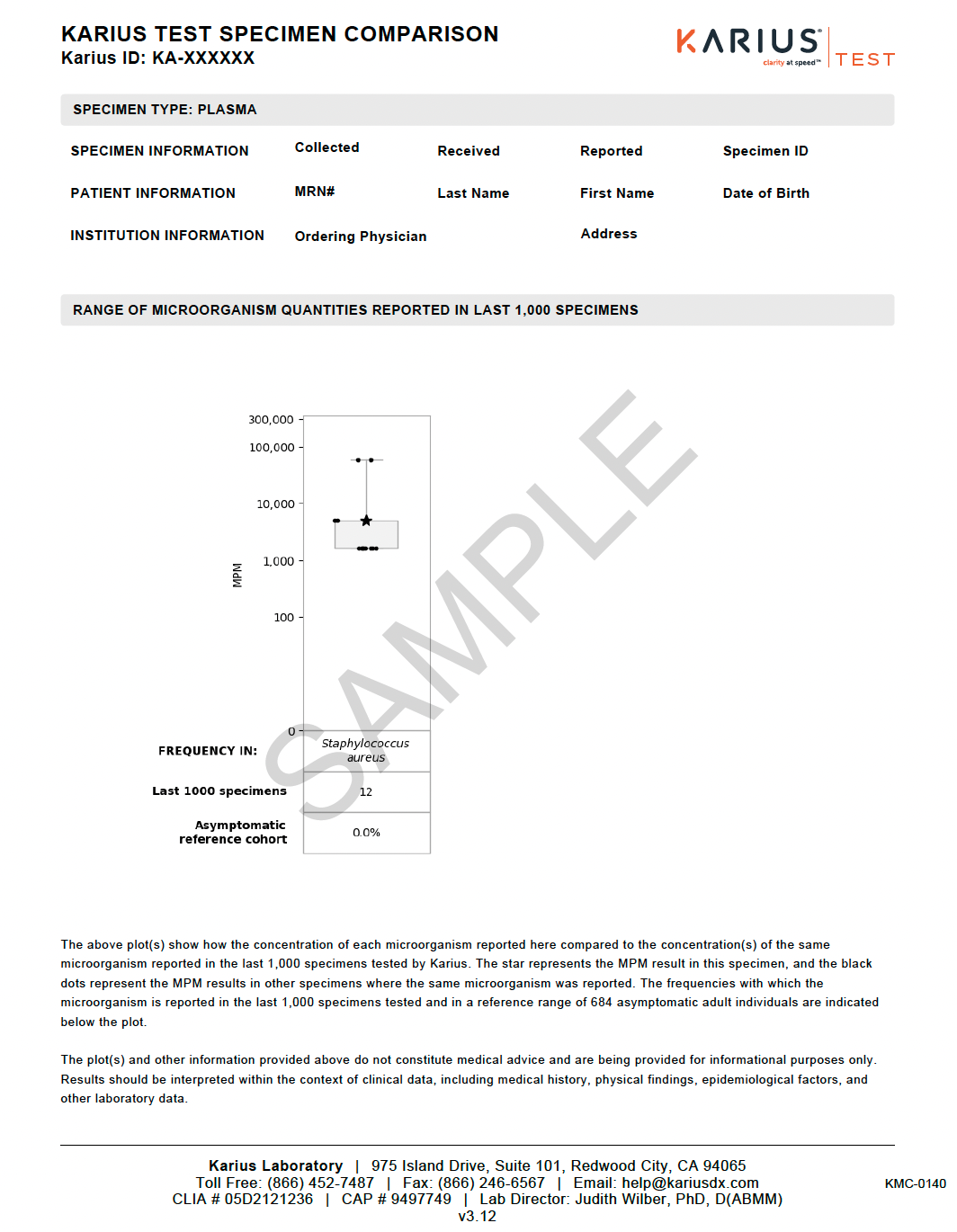

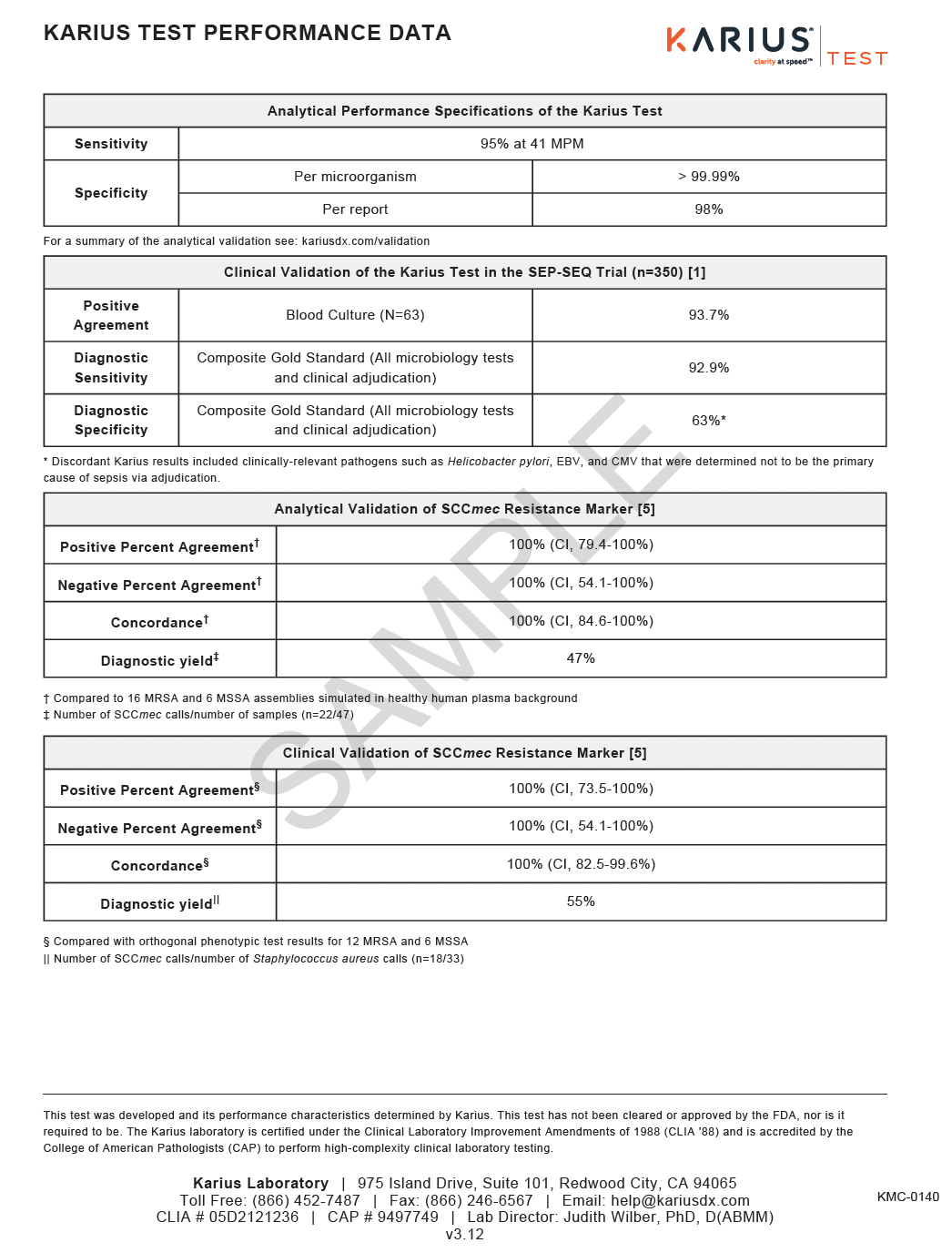

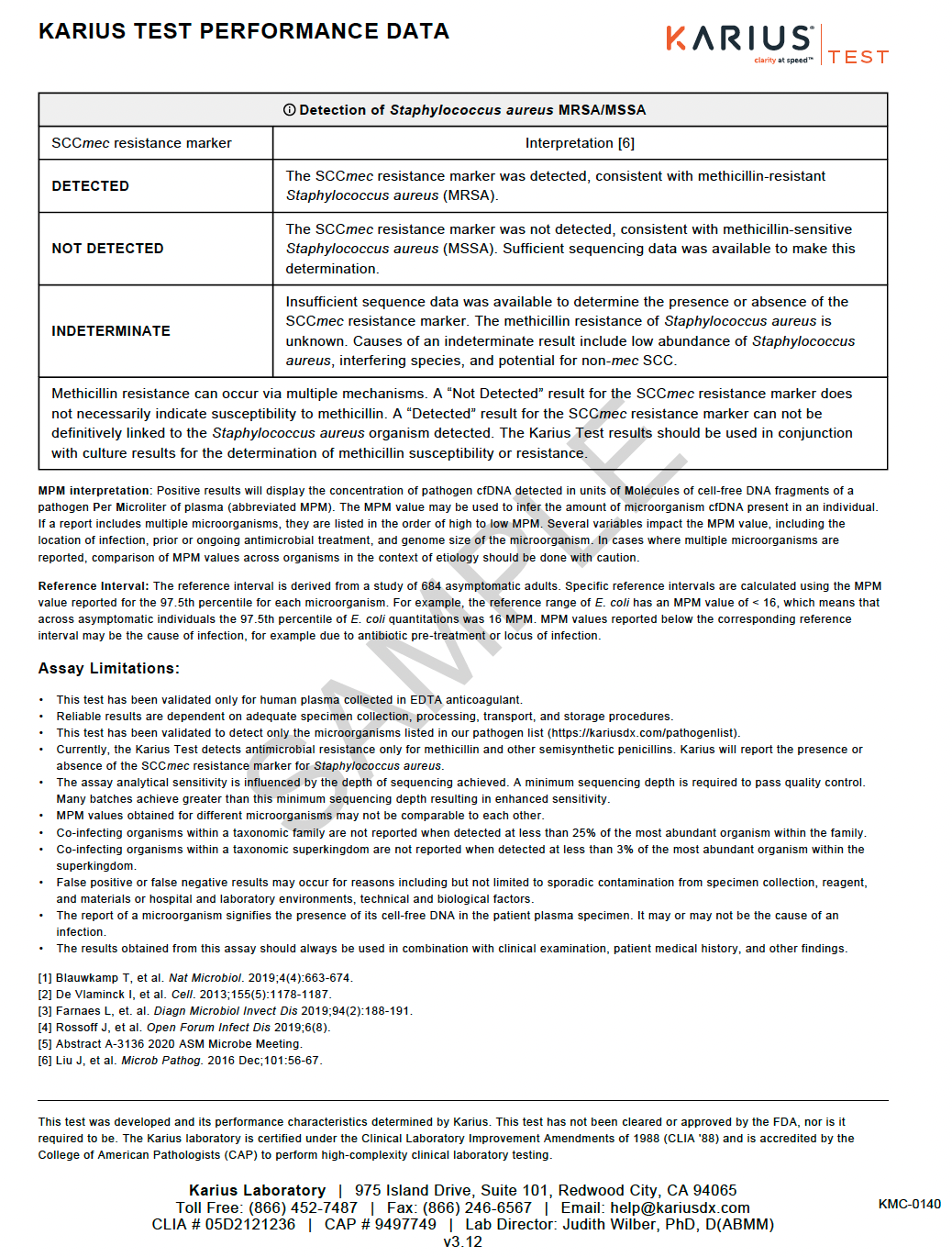
